# Lung function variability and the burden of COPD and lung restriction in South Africa and Burkina Faso: results from the AWI-Gen study

**DOI:** 10.64898/2026.09.22.26363668

**Authors:** Emmanuel Adonyo, Jing Chen, Catherine John, Richard Packer, Michele Ramsay, Chiara Batini, Richard van Zyl-Smit, Mervyn Mer, Palwende Romuald Boua, Godfred Agongo, F Xavier Gómez-Olivé, Reneilwe Given Mashaba, Martin D. Tobin, Anna L. Guyatt, Lisa K. Micklesfield

## Abstract

**Background:** Population-based spirometry data from sub-Saharan Africa remain limited. We characterised lung function, chronic obstructive pulmonary disease (COPD), lung restriction and associated factors from two Sub-Saharan African countries.

**Methods:** We performed spirometry using ndd Easy On-PC spirometers among adults aged ≥40 years from population-based cohorts in four communities in Burkina Faso and South Africa from January 2019–April 2022. We included 1,619 participants (936 women, 683 men) with repeatable forced expiratory volume in one second (FEV_1_) and forced vital capacity (FVC). COPD was defined as FEV_1_/FVC<0.70 and lung restriction as FEV_1_/FVC≥0.70, with FVC<80% predicted. We used multivariable models to test factors associated with lung function measurements (FEV_1_, FVC and FEV_1_/FVC), COPD and lung restriction.

**Results:** Median FEV_1_ and FVC ranged from 1.940–2.273L and 2.521–2.883L, respectively. COPD prevalence was 16.1% (95% CI 14.4–18.0; site range 10.8%–24.5%) and lung restriction prevalence was 21.3% (95% CI 19.4–23.4; site range 11.3%–36.2%). Prior tuberculosis was associated with higher odds of COPD (OR 2.86, 95% CI 1.75–4.59) and lower FEV_1_, FVC and FEV_1_/FVC. HIV infection was associated with higher COPD odds (OR 1.67, 95% CI 1.01–2.72) and lower FEV_1_/FVC. Hypertension was associated with higher lung restriction odds (OR 1.46, 95% CI 1.07–2.00) and lower FEV_1_ and FVC. Diabetes was associated with lower COPD odds (OR 0.50, 95% CI 0.24–0.94), lower FVC and higher FEV_1_/FVC. Ever-smoking and older age were associated with higher COPD odds (OR 2.21, 95% CI 1.48–3.32; OR 1.05, 95% CI 1.03–1.07, respectively); older age was associated with higher lung restriction odds (OR 1.02, 95% CI 1.00–1.04).

**Conclusion:** This multisite study highlights substantial heterogeneity in COPD and lung restriction prevalence across African populations and identifies respiratory multimorbidity with infectious and cardiometabolic conditions. These findings provide population-based evidence to inform respiratory surveillance in sub-Saharan Africa.

## Introduction

Chronic obstructive pulmonary disease (COPD) is a major global health challenge characterised by persistent airflow limitation resulting from airway and/or alveolar abnormalities, most commonly associated with long-term exposure to noxious particles or gases, particularly tobacco smoke(1,2). COPD caused approximately 3.4 million deaths globally in 2023(3), with a disproportionate burden in low- and middle-income countries (LMICs), including sub-Saharan Africa (SSA), where it is a major cause of morbidity and mortality(4–6). The burden of COPD in SSA is projected to rise steeply by 2050 due to population ageing, persistent infectious diseases, urbanisation, tobacco exposure and environmental risk factors(5–8). Despite its growing burden, COPD remains substantially under-recognised and underdiagnosed across SSA(5,6,9– 13), largely because of competing public health priorities, restricted access to spirometers, shortages of trained personnel, and other resource limitations(7,14,15). Spirometry provides objective measurements of airflow and lung volume, including forced expiratory volume in one second (FEV_1_), forced vital capacity (FVC), the FEV_1_/FVC ratio and peak expiratory flow (PEF)(16,17). These measures are used to diagnose and monitor chronic respiratory diseases, particularly COPD and asthma, and are established predictors of morbidity and mortality across the life course(18,19).

The Burden of Obstructive Lung Disease (BOLD) study, an international population-based spirometry study, reported substantial variation in the prevalence of COPD and lung restriction across its African sites(20,21). Across African BOLD sites, COPD prevalences ranged from 5.6% in Gezira, Sudan, to 18.9% in Cape Town, South Africa, while a subsequent BOLD analysis reported site-specific prevalences ranging from 6.9% in Blantyre, Malawi, to 19.3% in Uitsig/Ravensmead, Cape Town, South Africa(22–24). A meta-analysis incorporating studies with heterogeneous designs and case definitions reported COPD prevalences ranging from 1.7% in Kampala, Uganda, to 24.8% in Cape Town, South Africa(5). Studies on lung restriction in African populations have suggested that it may be more common than COPD in some settings(25,26). However, there are few harmonised multisite African studies that jointly characterise lung function, COPD and lung restriction and examine demographic, socioeconomic, behavioural, clinical and environmental factors critical to understanding disease burden.

Our study contributes to addressing this gap by conducting standardised spirometry across three sites in South Africa (Agincourt, **Di**kgale, **Ma**mabolo and **Mo**thiba (DIMAMO) and Soweto) and one site in Burkina Faso (Nanoro) within the framework of the Africa Wits-INDEPTH Partnership for Genomic Studies (AWI-Gen)(27,28). We applied rigorous training and quality control, characterised lung function measures, estimated the prevalence of COPD and lung restriction, and examined associations with demographic, socioeconomic, behavioural, clinical and environmental factors. We then compared our findings with relevant regional studies to strengthen the epidemiological utility of spirometry for understanding respiratory health in sub-Saharan Africa.

## Methods

### AWI-Gen study design and participants

The AWI-Gen study is a population-based cohort study conducted across six sites in four African countries: Kenya, Ghana, Burkina Faso and South Africa(28). The AWI-Gen cohort initially recruited 12,032 adults aged ≥40 years during Wave 1 (2011–2015) across the six sites. During Wave 2 (2019–2022), data were collected from 7,807 participants. Exclusion criteria for AWI-Gen included first-degree relatives of enrolled participants, pregnant women, recent immigrants and individuals with physical impairments precluding data collection.

Spirometry measurements were collected as part of the AWI-Gen XHALE substudy conducted during Wave 2 at four sites: Nanoro in Burkina Faso, and DIMAMO, Agincourt and Soweto in South Africa (Figure 1). Trained researchers administered a respiratory health questionnaire before spirometry. Individuals with medical contraindications to spirometry according to ATS/ERS recommendations were excluded(17,19), including recent myocardial infarction, uncontrolled cardiovascular instability, recent thoracic, abdominal or ocular surgery, pneumothorax, and active respiratory infection.

**Figure 1.**
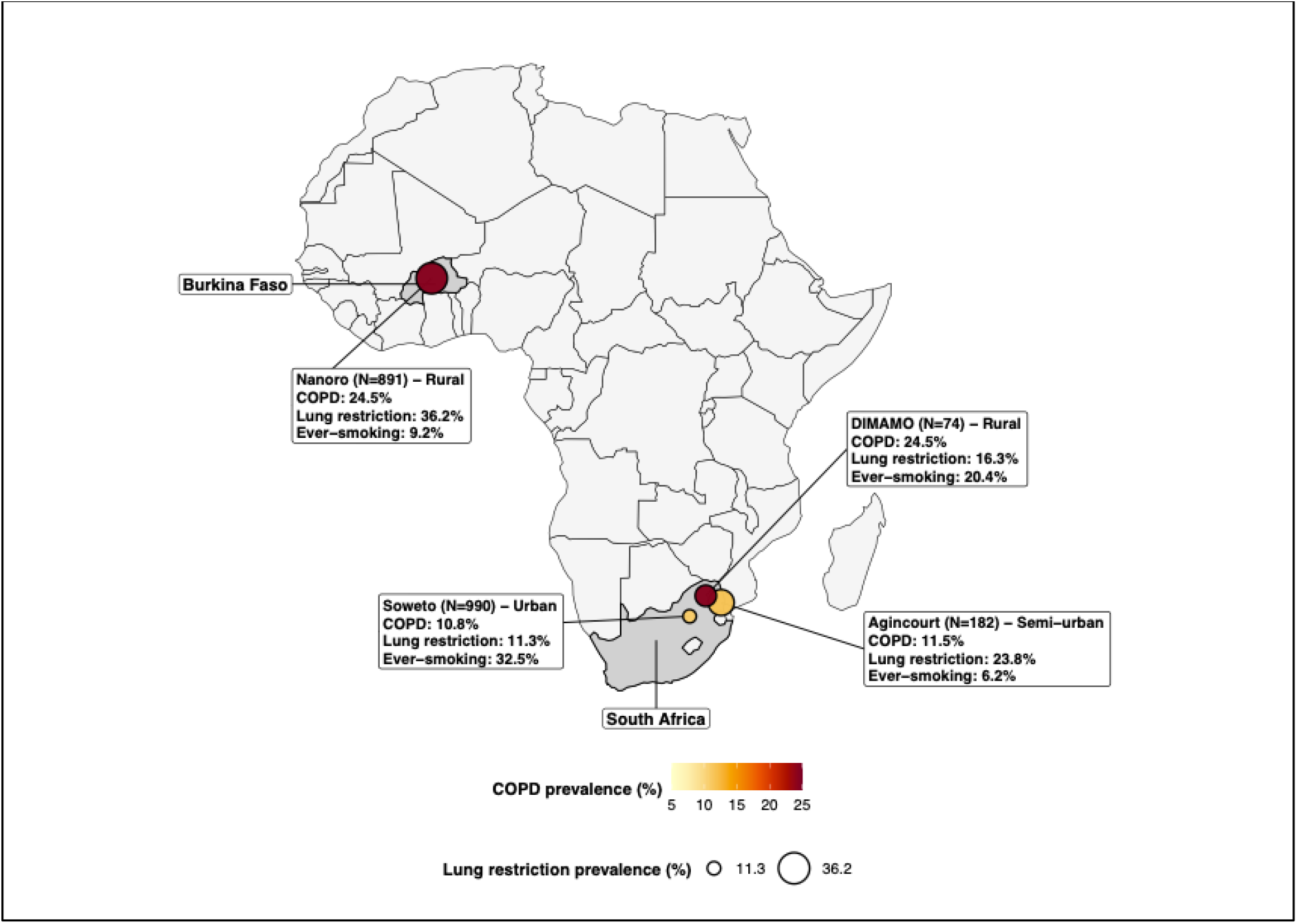
Geographic distribution of the four AWI-Gen study sites included in this analysis. Site locations are shown with coloured dots; labels give the site-specific numbers of participants with FVC measurements available before quality control.

The study was approved by the Human Research Ethics Committee (Medical) of the University of the Witwatersrand (certificate numbers M121029, M170880 and M2210108). Each participating study site obtained local ethics approval in accordance with institutional and national requirements. All participants provided written informed consent. Demographic, anthropometric, lifestyle, clinical and environmental data were collected before spirometry.

### Spirometry procedures

Spirometry testing was conducted using ndd Easy On-PC spirometers, which were factory-calibrated and verified prior to data collection using a 3-litre syringe within a ±3% tolerance(19). Participants completed a minimum of three valid blows, with up to eight attempts permitted.

Spirometry measurements included forced expiratory volume in one second (FEV_1_; L), forced vital capacity (FVC; L), the FEV_1_/FVC ratio and peak expiratory flow (PEF; L/min). Where bronchodilator responsive testing was completed, participants with an initial FEV_1_/FVC <0.70 received 400 µg of salbutamol via a spacer and were retested after 10–20 minutes to assess bronchodilator responsiveness.Those with persistent post-bronchodilator obstruction were referred for clinical follow-up. Spirometry data were stored electronically, backed up daily and exported weekly to a Research Electronic Data Capture (REDCap) repository(29).

### Quality control

Participants were eligible for inclusion in this analysis if they produced at least two successful spirometry manoeuvres. For each participant, the “best” FEV_1_ and “best” FVC were defined as the highest values obtained across acceptable manoeuvres and were not required to come from the same manoeuvre. The FEV_1_/FVC ratio was calculated from these selected best FEV_1_ and FVC values. Repeatability was assessed as previously described(30): the difference between the best FEV_1_ and any other blow was required to be ≤250 mL, with the same criterion applied to FVC. Participants with repeatable FEV_1_ and FVC were retained for analysis. We conducted quality control separately for pre- and post-bronchodilator blows at each study site to identify site-level variation in performance.

### Definition of COPD and lung restriction

Predicted values for FEV_1_, FVC and FEV_1_/FVC were calculated using National Health and Nutrition Examination Survey III (NHANES III) equations for African American adults(31), from which percentage-predicted values were derived. NHANES III equations were used because no spirometric reference equation has been validated across the continental African populations included in this study, and recent African evidence has questioned the suitability of the recent GLI-2022 race-neutral equations in settings where the underpinning reference dataset does not include prospectively collected healthy data from sub-Saharan African countries(32). Because post-bronchodilator spirometry was available only for a small subset of participants, COPD was defined using pre-bronchodilator spirometric evidence of obstruction, based on FEV_1_/FVC<0.70 (GOLD 1–4)(1,33), and lung restriction as FEV_1_/FVC ≥0.70 with FVC <80% predicted(34,35).

Prevalence estimates for COPD and lung restriction were calculated overall and by study site.

### Covariates and exposures

Covariates included age, height, sex (female as the reference), smoking status (ever-smoking, defined as ≥100 cigarettes smoked during a lifetime; never-smoking as the reference), study site (Soweto as the reference), and socioeconomic status (SES). For this analysis, SES was calculated as the proportion of household assets owned by a participant, normalised by the total assets assessed at the participant’s study site, and grouped into quintiles, with higher quintiles indicating higher household asset ownership, and modelled as a continuous variable(36). Exposures included “Solid fuel exposure”, which was categorised as “solid fuel use with no kitchen ventilation”, “solid fuel use with kitchen ventilation”, or “no solid fuel use” (reference). Self-reported, clinician-diagnosed conditions included prior tuberculosis (TB), chronic kidney disease (CKD), diabetes, hypertension, and cardiovascular disease (CVD), where CVD was defined as any of stroke, transient ischaemic attack, angina, myocardial infarction or heart failure). Human immunodeficiency virus (HIV) testing was optional in South Africa and was not offered in Burkina Faso, where national prevalence was >2%(37).

### Assessment of potential selection bias

To assess potential selection bias, we compared age, sex, height, weight, tobacco smoking and SES, measured at AWI-Gen wave 1, across four groups: (i) participants from the four study sites with available tobacco smoking data at AWI-Gen wave 1; (ii) participants from group (i) with AWI-Gen wave 2 data; (iii) participants who completed the respiratory health questionnaire (RHQ) in AWI-Gen wave 2; and (iv) participants with repeatable spirometry in AWI-Gen wave 2. These groups were defined solely for comparing AWI-Gen wave 1 characteristics and do not represent consecutive stages in the selection of the analytical population. Participants newly recruited in DIMAMO during AWI-Gen wave 2 had no AWI-Gen wave 1 data and therefore could not contribute to this comparison; however, they remained eligible for inclusion in the study. Continuous variables were summarised using medians and interquartile ranges, and categorical variables using counts and percentages.

### Statistical analysis

Univariable linear regression was used to estimate crude associations between each exposure and FEV_1_, FVC and FEV_1_/FVC as a face validity check to confirm expected associations with established determinants of lung function (Supplementary Table S2). Univariable logistic regression was used to estimate crude associations with COPD and lung restriction to provide baseline estimates before adjustment (Supplementary Tables S3 and S4). Selection of variables for the multivariable models was based on prior literature and was not informed by the univariable analyses. For the main analysis, multivariable logistic regression models estimated adjusted associations with COPD and lung restriction (binary outcomes), and multivariable linear regression models estimated adjusted associations with FEV_1_, FVC and FEV_1_/FVC (continuous outcomes). For each outcome, the base model included age, height, sex, smoking status, study site and SES. Age and height were modelled as continuous variables; age was reported per year, height per metre in the linear models but rescaled to 10 cm increments in the logistic models to avoid odds ratios per one-metre height difference, and SES quintile was modelled as a continuous score. Each key exposure, comprising solid fuel exposure, TB, diabetes, CVD, hypertension, CKD or HIV, was then added separately to the base model. Each model included participants with complete data for the outcome, base model covariates and the additional exposure; sample sizes therefore varied across exposure-specific models.

Adjusted estimates from logistic regression are presented as ORs with 95% CIs, and estimates from linear regression as β coefficients with 95% CIs. The proportion of variance explained by each linear regression model was summarised using R^2^. No formal correction for multiple comparisons was applied; estimates were interpreted in the context of the prespecified exposures and multiple exposure-specific models. Analyses were conducted using R (R Foundation for Statistical Computing, Vienna, Austria; https://www.r-project.org/).

## Results

### Assessment of potential selection bias

Table 1 compares characteristics measured at AWI-Gen wave 1 across the four comparison groups. Participants with repeatable spirometry were younger than AWI-Gen 1, with a median age of 50 years compared with 51–52 years in the other groups, had the highest proportion in the highest SES quintile (26.0% compared with 20.0%–22.5% in other groups), and were more likely to be women (57.6%). Participants who completed the respiratory health questionnaire had the lowest proportion of ever smokers (19.1%). Height and weight were similar across the groups.

**Table 1.** AWI-Gen wave 1 characteristics in the four-site AWI-Gen population and respiratory-study comparison groups.

|  | **AWI-Gen 1**<br>**N = 8,004** | **AWI-Gen 2**<br>**N = 4,825** | **Completed RHQ**<br>**N = 3,496** | **Repeatable spirometry**<br>**N = 1,618** |
| --- | --- | --- | --- | --- |
| Age (years) | 52 (46, 58) | 51 (45, 57) | 52 (46, 57) | 50 (45, 55) |
| Sex (Female) | 4,450 (55.6%) | 2,742 (56.8%) | 2,005 (57.4%) | 932 (57.6%) |
| Height (m) | 1.64 (1.58, 1.71) | 1.64 (1.59, 1.71) | 1.65 (1.59, 1.72) | 1.65 (1.59, 1.71) |
| Weight (kg) | 68 (56, 81) | 68 (57, 82) | 66 (55, 79) | 69 (57, 84) |
| Ever-smoking | 2,049 (25.6%) | 1,146 (23.8%) | 666 (19.1%) | 396 (24.5%) |
| SES quintile |  |  |  |  |
| 1 (lowest) | 1,601 (20.0%) | 843 (17.5%) | 670 (19.2%) | 233 (14.4%) |
| 2 | 1,601 (20.0%) | 915 (19.0%) | 714 (20.4%) | 293 (18.1%) |
| 3 | 1,601 (20.0%) | 939 (19.5%) | 695 (19.9%) | 309 (19.1%) |
| 4 | 1,601 (20.0%) | 1,040 (21.6%) | 697 (19.9%) | 363 (22.4%) |
| 5 (highest) | 1,600 (20.0%) | 1,088 (22.5%) | 720 (20.6%) | 420 (26.0%) |
| *Data are n (%) or median (25th, 75th percentile). RHQ, respiratory health questionnaire; SES, socioeconomic status. All characteristics were measured at AWI-Gen wave 1. Each later column includes only participants with AWI-Gen wave 1 data who were also present in the corresponding later study group. The table assesses potential selection bias and does not describe selection of the analytical population. The repeatable-spirometry column includes 1,618 of the 1,619 final analytical participants because one participant lacked AWI-Gen wave 1 data.* |  |  |  |  |

### Spirometry quality control

Figure 1 shows geographic distribution of the AWI-Gen spirometry study sites, while participant selection and exclusions during spirometry quality control are summarised in Table 2. Following exclusion of tests without FVC measurements(38), spirometry data were available for 2,137 participants. Repeatable FEV_1_ was obtained for 1,776 participants (83.1%) and repeatable FVC for 1,664 (77.9%). Overall, 1,629 participants (76.2%) achieved both repeatable FEV_1_ and FVC; the proportion was highest in Soweto (871/990; 88.0%) and lowest in Nanoro (578/891; 64.9%). Ten participants with repeatable spirometry lacked smoking data and were excluded, giving a final analytical sample of 1,619 participants: 130 from Agincourt, 49 from DIMAMO, 575 from Nanoro and 865 from Soweto.

**Table 2.** AWI-Gen participant selection process to identify analytical samples with repeatable spirometry. We undertook quality control per study site.

| **Selection process** | **Overall**<br>**N = 2,137** | **Agincourt**<br>**N = 182** | **DIMAMO**<br>**N = 74** | **Nanoro**<br>**N = 891** | **Soweto**<br>**N = 990** |
| --- | --- | --- | --- | --- | --- |
| At least two blows | 1,858 (86.9%) | 150 (82.4%) | 54 (73.0%) | 723 (81.1%) | 931 (94.0%) |
| At least one acceptable blow | 1,858 (86.9%) | 150 (82.4%) | 54 (73.0%) | 723 (81.1%) | 931 (94.0%) |
| Repeatable FEV<sub>1</sub> | 1,776 (83.1%) | 145 (79.7%) | 51 (68.9%) | 666 (74.7%) | 914 (92.3%) |
| Repeatable FVC | 1,664 (77.9%) | 133 (73.1%) | 50 (67.6%) | 602 (67.6%) | 879 (88.8%) |
| Repeatable FEV<sub>1</sub> and FVC | 1,629 (76.2%) | 131 (72.0%) | 49 (66.2%) | 578 (64.9%) | 871 (88.0%) |
| Missing smoking data | 10 (0.5%) | 1 (0.5%) | 0 (0.0%) | 3 (0.3%) | 6 (0.6%) |
| Final analytical sample | 1,619 (75.8%) | 130 (71.4%) | 49 (66.2%) | 575 (64.5%) | 865 (87.4%) |

### Demographics and lung function characteristics

Characteristics of the study population are summarised in Table 3. Overall, 57.8% of participants were female and 21.7% reported ever-smoking, with the highest prevalence in Soweto (32.5%). Median age was 55 years (IQR 50–60); Agincourt had the highest median age (62 years; IQR 54–68).

**Table 3.** Demographic and lung function characteristics of the AWI-Gen analytical population.

|  | **Overall**<br>**N = 1,619** | **Agincourt**<br>**N = 130** | **DIMAMO**<br>**N = 49** | **Nanoro**<br>**N = 575** | **Soweto**<br>**N = 865** |
| --- | --- | --- | --- | --- | --- |
| Sex (Female) | 936<br>(57.8%) | 86<br>(66.2%) | 30<br>(61.2%) | 338<br>(58.8%) | 482<br>(55.7%) |
| Ever-smoking | 352<br>(21.7%) | 8<br>(6.2%) | 10<br>(20.4%) | 53<br>(9.2%) | 281<br>(32.5%) |
| Age (years) | 55<br>(50, 60) | 62<br>(54, 68) | 59<br>(53, 62) | 54<br>(50, 59) | 55<br>(50, 60) |
| Height (metres) | 1.64<br>(1.58, 1.71) | 1.63<br>(1.59, 1.69) | 1.61<br>(1.55, 1.68) | 1.65<br>(1.60, 1.72) | 1.63<br>(1.57, 1.71) |
| Weight (kg) | 69.1<br>(56.8, 84.1) | 75.1<br>(63.0, 87.0) | 68.0<br>(58.7, 86.1) | 56.8<br>(50.4, 65.0) | 78.7<br>(66.4, 92.7) |
| FEV<sub>1</sub> (L) | 2.114<br>(1.726, 2.557) | 1.979<br>(1.623, 2.272) | 2.014<br>(1.589, 2.425) | 1.940<br>(1.532, 2.329) | 2.273<br>(1.920, 2.769) |
| FVC (L) | 2.711<br>(2.262, 3.332) | 2.524<br>(2.141, 3.033) | 2.573<br>(2.140, 3.332) | 2.521<br>(2.105, 3.064) | 2.883<br>(2.399, 3.519) |
| FEV<sub>1</sub>/FVC | 0.790<br>(0.735, 0.832) | 0.774<br>(0.736, 0.808) | 0.763<br>(0.709, 0.807) | 0.769<br>(0.702, 0.830) | 0.800<br>(0.755, 0.836) |
| % Predicted FEV<sub>1</sub> | 89.6<br>(75.6, 102.4) | 89.1<br>(74.7, 100.0) | 91.4<br>(71.7, 108.7) | 78.5<br>(66.5, 89.3) | 97.4<br>(85.8, 108.8) |
| % Predicted FVC | 91.1<br>(78.9, 102.7) | 90.4<br>(77.7, 101.0) | 97.1<br>(78.1, 109.7) | 82.1<br>(71.6, 92.7) | 97.3<br>(86.1, 108.2) |
| COPD<br>(GOLD 1–4) | 261<br>(16.1%) | 15<br>(11.5%) | 12<br>(24.5%) | 141<br>(24.5%) | 93<br>(10.8%) |
| Moderate-to-severe<br>obstruction (GOLD 2–4) | 194<br>(12.0%) | 11<br>(8.5%) | 8<br>(16.3%) | 117<br>(20.3%) | 58<br>(6.7%) |
| Lung<br>restriction | 345<br>(21.3%) | 31<br>(23.8%) | 8<br>(16.3%) | 208<br>(36.2%) | 98<br>(11.3%) |
| *Data are n (%) or median (25th, 75th percentile). COPD, chronic obstructive pulmonary disease; FEV<sub>1</sub>, forced expiratory volume in one second; FVC, forced vital capacity; GOLD, Global Initiative for Chronic Obstructive Lung Disease. Percentage-predicted values were derived using NHANES III equations for African American adults. COPD categories are based on pre-bronchodilator spirometry; lung restriction was defined as FEV<sub>1</sub>/FVC ≥0.70 with FVC <80% predicted. Formal between-site statistical comparisons are not shown in this descriptive table.* |  |  |  |  |  |

Median FEV_1_ ranged from 1.940 L in Nanoro to 2.273 L in Soweto, and median FVC ranged from 2.521 L in Nanoro to 2.883 L in Soweto. Median percentage-predicted FEV_1_ ranged from 78.5% in Nanoro to 97.4% in Soweto, and median percentage-predicted FVC ranged from 82.1% in Nanoro to 97.3% in Soweto. Median FEV_1_/FVC was 0.790 overall, ranging from 0.763 in DIMAMO to 0.800 in Soweto.

### Prevalence of COPD and lung restriction

Overall, COPD prevalence was 16.1%, highest in Nanoro and DIMAMO (both 24.5%) and lowest in Soweto (10.8%) (Table 2). Moderate-to-severe obstruction (GOLD 2–4) prevalence was 12.0%, again highest in Nanoro (20.3%) and lowest in Soweto (6.7%). Lung restriction prevalence was 21.3%, ranging from 11.3% in Soweto to 36.2% in Nanoro.

Among the 261 participants with pre-bronchodilator COPD (GOLD 1–4), 38 (14.6%) underwent bronchodilator responsiveness testing to assess whether obstruction persisted after bronchodilation. Of these, 21 (55.3%) had persistent obstruction (FEV_1_/FVC<0.70).

### Factors associated with lung function, COPD and lung restriction

Univariable regression analyses identified crude associations between several demographic, behavioural, clinical and environmental factors and the lung function, COPD and lung restriction outcomes (Supplementary Tables S2–S4).

Full adjusted estimates and 95% CIs for the multivariable linear regression models are presented in Supplementary Tables S5–S7. Age, height, sex, smoking status, study site and SES together explained 52.0% of the variance in FEV_1_, 56.9% of the variance in FVC and 6.0% of the variance in FEV_1_/FVC, based on the model R^2^ values. Older age, shorter height and female sex were associated with lower FEV_1_ and FVC, while older age and ever-smoking were associated with lower FEV_1_/FVC. Ever-smoking was associated with lower FEV_1_ and FEV_1_/FVC, but there was no evidence of an association with FVC after adjustment. Higher SES was associated with higher FEV_1_ and FVC, but not with FEV_1_/FVC. After adjustment for base-model covariates, prior TB and hypertension were associated with lower FEV_1_ and FVC, while diabetes was associated with lower FVC but not FEV_1_. Prior TB and HIV infection were associated with lower FEV_1_/FVC, whereas diabetes and CVD were associated with higher FEV_1_/FVC. Solid fuel exposure and CKD were not independently associated with FEV_1_, FVC or FEV_1_/FVC.

Multivariable logistic regression analyses identifying correlates of COPD and lung restriction are presented in Figure 2 and Figure 3, respectively, with full model estimates presented in Supplementary Tables S8 and S9. Ever smokers had approximately twofold higher odds of COPD than never smokers, while each additional year of age was associated with 5% to 6% higher odds of COPD. Among clinical predictors, prior TB (OR 2.86, 95% CI 1.75–4.59) and HIV infection (OR 1.67, 95% CI 1.01–2.72) were associated with higher odds of COPD, whereas diabetes was associated with 50% lower odds (OR 0.50, 95% CI 0.24–0.94). Although male sex and solid fuel exposure were associated with higher odds of COPD in the univariable models, neither remained independently associated with COPD after adjustment.

**Figure 2.**
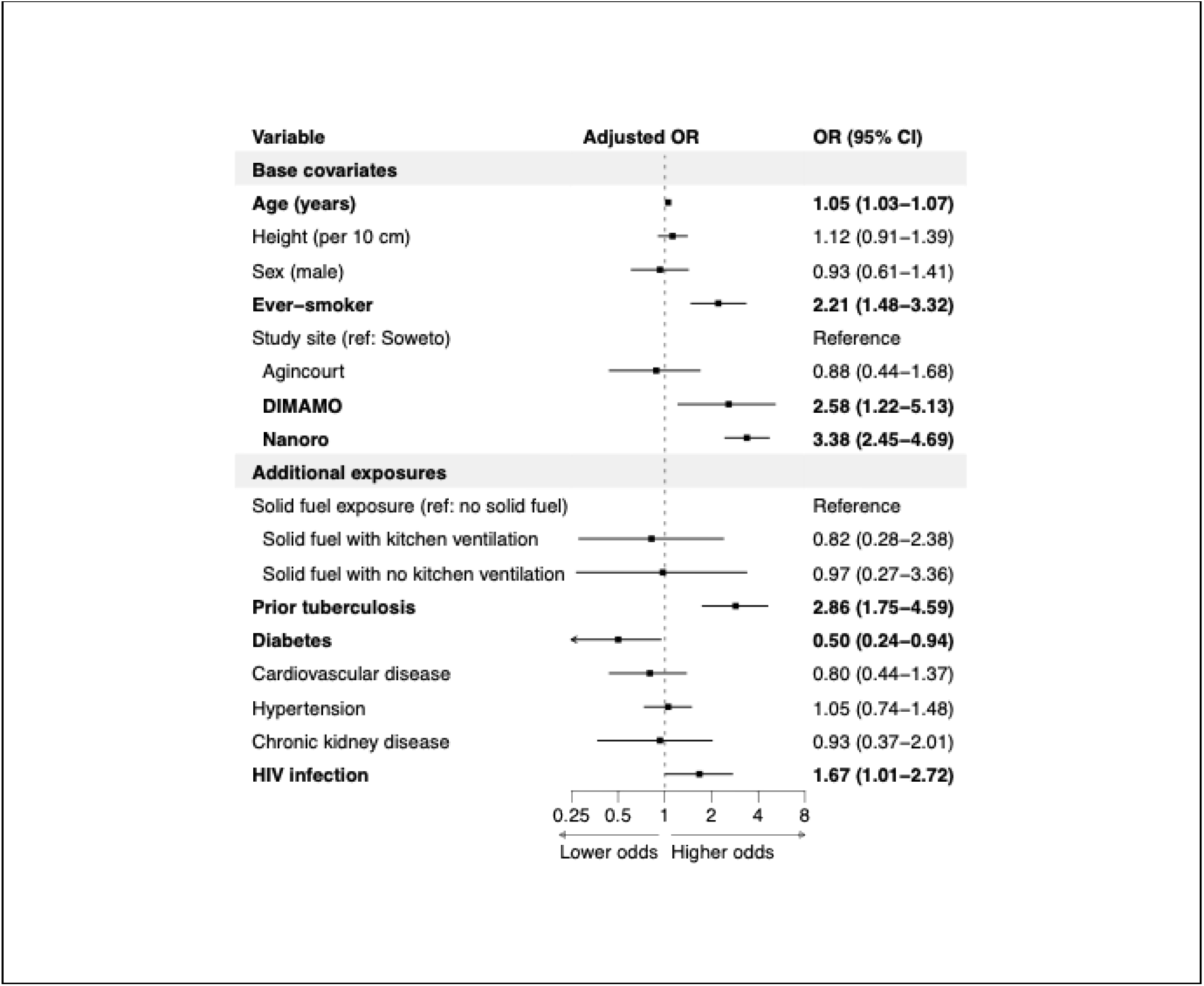
Multivariable logistic regression analyses of COPD in the AWI-Gen analytical population. Note: Values are adjusted ORs (95% CIs). Base models included age, height, sex, ever-smoking, study site and SES (continuous quintile score); each additional exposure was modelled separately, so N varies with data completeness. TB, tuberculosis; CVD, cardiovascular disease; CKD, chronic kidney disease; HIV, human immunodeficiency virus.

**Figure 3.**
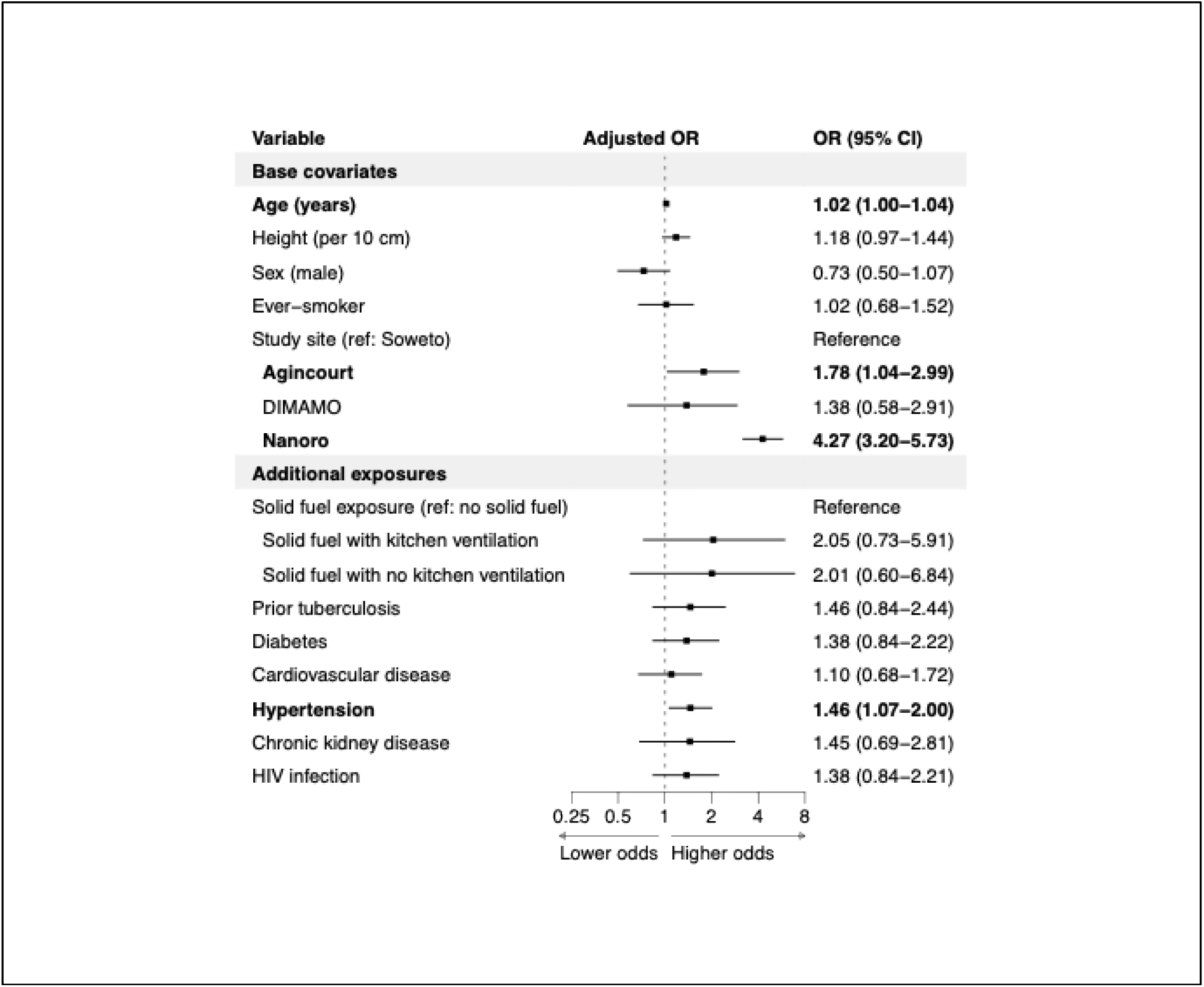
Multivariable logistic regression analyses of lung restriction in the AWI-Gen analytical population. Note: Values are adjusted ORs (95% CIs). Base models included age, height, sex, ever-smoking, study site and SES (continuous quintile score); each additional exposure was modelled separately, so N varies with data completeness. TB, tuberculosis; CVD, cardiovascular disease; CKD, chronic kidney disease; HIV, human immunodeficiency virus.

For lung restriction, each additional year of age was associated with 2% to 4% higher odds of lung restriction. Hypertension was the only clinical condition associated with lung restriction after adjustment (OR 1.46, 95% CI 1.07–2.00). Although ever-smoking and higher SES were associated with lower odds of lung restriction and solid fuel exposure was associated with higher odds in the univariable models, none of these factors remained independently associated with lung restriction after adjustment.

## Discussion

In this multi-site population-based study of 1,619 adults across four communities in Burkina Faso and South Africa, we observed substantial variation in lung function and respiratory impairment across study sites. The prevalences of both COPD and lung restriction were lowest in Soweto, South Africa (10.8% and 11.3%, respectively), while COPD was highest in Nanoro, Burkina Faso, and DIMAMO, South Africa (24.5% each), and lung restriction was highest in Nanoro (36.2%). We have shown that respiratory impairment co-existed with other chronic diseases, including HIV infection and hypertension, while prior tuberculosis, older age and ever-smoking were confirmed as risk factors for COPD. Diabetes was associated with lower FVC, higher FEV_1_/FVC and lower odds of COPD.

Reviews from previous studies have reported a high burden of COPD and lung restriction in Africa, although estimates vary widely due to differences in study design, spirometry procedures, outcome definitions, reference equations and quality control processes(5,6). The AWI-Gen COPD prevalence of 16.1% was higher than estimates reported from several BOLD African sites, including Gezira, Sudan (5.6%); Blantyre, Malawi (8.2%); and Ile-Ife, Nigeria (7.7%), but was closer to estimates from Chikhwawa, Malawi (14.0%) and Cape Town, South Africa (18.9%)(5,23,39). Several factors may explain these differences. BOLD COPD prevalence estimates were based on post-bronchodilator FEV_1_/FVC below the lower limit of normal (LLN)(40), defined as the lower 5th percentile of the predicted FEV_1_/FVC distribution in healthy never-smoking reference populations of similar age, sex, height, and ancestry or ethnic group, whereas our study defined COPD using pre-bronchodilator FEV_1_/FVC <0.70. In addition, use of a fixed FEV_1_/FVC threshold may overestimate COPD in older adults because FEV_1_/FVC declines with age, particularly in mild disease(41,42). BOLD African participants were slightly younger than our participants, with mean ages of 50.5–53.4 years compared with 55.4 years in our study.

Within our study, COPD prevalence also varied substantially across urban and rural settings in West and South Africa. The lowest prevalence was observed in urban Soweto, South Africa (10.8%), where the prevalence of ever-smoking was highest (32.5%), whereas the highest prevalences were observed in rural DIMAMO, South Africa (24.5%), and rural Nanoro, Burkina Faso (24.5%), where the prevalence of ever-smoking was lower (20.4% and 9.2%, respectively). This pattern should not be interpreted as an inverse association between tobacco smoking and COPD because ever-smoking was associated with higher odds of COPD in both the univariable and multivariable models, with more than twofold higher odds after adjustment. Approximately 20%–30% of COPD cases occur among never-smokers, and this proportion may be higher in resource-limited settings(43,44). The between-site variation in our study therefore suggests that the COPD burden may reflect different combinations of tobacco and non-tobacco exposures. In rural settings, these may include household air pollution, childhood exposure to environmental tobacco smoke, previous respiratory infections, asthma, occupational exposures, socioeconomic disadvantage and early-life adversity(43,45). Although self-reported solid-fuel exposure was associated with COPD and lung restriction in the univariable models, neither association remained after adjustment for the base-model covariates, and the adjusted estimates were imprecise. This may reflect exposure misclassification because current cooking fuel does not capture cumulative household air pollution, ventilation quality, time spent near cooking areas, previous fuel use or occupational inhalational exposures(45).

Lung restriction was also common in our study, with an overall prevalence of 21.3%. This was slightly higher than, but broadly comparable to, estimates from Yaoundé, Cameroon, where lung restriction prevalence was 18.8% using FEV_1_/FVC ≥LLN with FVC <LLN and 15.0% using FVC

<80% predicted with FEV_1_/FVC ≥LLN, illustrating how restriction estimates depend on the definition of low FVC(25). However, our estimate was lower than those reported from African BOLD sites, including Cotonou, Benin (78.4%); Ile-Ife, Nigeria (71.5%); Cape Town, South Africa (46.8%); and Blantyre, Malawi (48.0%)(26). These estimates are also sensitive to the reference equations applied. In Ile-Ife, the prevalence of low FVC differed substantially when NHANES-III “Caucasian”, NHANES-III African American, Global Lung Function Initiative 2012 (GLI-2012) and locally derived equations were applied to the same population, with NHANES-III Caucasian equations giving higher estimates than the other equations(46). This may partly explain why our estimate, based on NHANES-III African American reference equations, was lower than BOLD African estimates derived using NHANES-III Caucasian equations(26,46). Within our study, lung restriction was highest in Nanoro, where participants had lower FVC, lower percent-predicted FVC and lower median body weight than participants in Soweto. The lower prevalence in DIMAMO than Nanoro, despite both being rural sites, indicates that rural residence alone does not explain the observed variation in low FVC in our study. Variation in lung restriction may therefore reflect site-specific combinations of nutritional, infectious, cardiometabolic, environmental and early-life factors, some of which were not fully captured in our analysis(25,26).

Respiratory impairments were associated with both cardiometabolic and infectious disease comorbidities. Diabetes was associated with lower odds of COPD as well as lower FVC and higher FEV_1_/FVC. Lower FVC among people with diabetes or impaired glucose metabolism has been reported in the Atherosclerosis Risk in Communities (ARIC) study, the Jackson Heart Study and COPDGene, including populations of African ancestry(47–49). Chronic hyperglycaemia may affect the lung compliance through connective tissue glycation, pulmonary microangiopathy, systemic inflammation and autonomic neuropathy(50,51). Unlike ARIC and a previous meta-analysis, which also reported lower FEV_1_, we did not find an association with FEV_1_, possibly reflecting differences in diabetes severity, disease duration, treatment patterns, smoking exposure or statistical power(47,52). The lower odds of COPD among participants with diabetes should not be interpreted as evidence that diabetes is protective. Because diabetes was associated with lower FVC but not FEV_1_, the resulting higher FEV_1_/FVC may have reduced the probability of meeting the ratio-based COPD definition. COPDGene similarly reported lower FVC and higher FEV_1_/FVC among participants with diabetes (49,53). Hypertension was associated with 46% higher odds of lung restriction as well as lower FEV_1_ and FVC, with FEV_1_ association only emerging after adjustment. Similar associations have been reported among hypertensive adults in Ethiopia, African Americans in the Jackson Heart Study and participants in the international BOLD study(26,54,55). These associations may reflect shared vascular, inflammatory and cardiac pathways underlying hypertension and reduced lung function(54). Ever-smoking, SES and solid-fuel exposure were not independently associated with lung restriction, CVD and CKD were not associated with lung restriction in either the multivariable or univariable analysis. These null findings indicate an absence of evidence rather than evidence that these exposures are irrelevant.

Prior tuberculosis and HIV infection were associated with higher odds of COPD and reduced lung function, but were not associated with lung restriction. HIV-associated abnormal spirometry has previously been reported in African cohorts(56,57), and pulmonary tuberculosis remains an important contributor to impaired lung function among people living with HIV(56– 58). Post-tuberculosis lung disease is characterised by heterogeneous airway and parenchymal abnormalities, including fibrosis, bronchiectasis, airway distortion, small-airway disease and parenchymal scarring, which may result in reduced lung volumes, airflow obstruction or mixed impairment(59,60). Although BOLD and a recent systematic review reported associations between prior tuberculosis and both airflow obstruction and lung restriction, we did not observe an association with lung restriction(60,61). This may reflect limited characterisation of post-tuberculosis lung damage using self-reported tuberculosis history, which did not distinguish recurrent disease, extent of parenchymal involvement, time since treatment or residual radiographic abnormalities(62).

Ever-smoking was associated with higher odds of COPD and with lower FEV_1_ and FEV_1_/FVC, consistent with evidence from sub-Saharan Africa and globally(5,24). Tobacco smoke promotes chronic airway inflammation, oxidative stress, mucus hypersecretion, airway remodelling and alveolar destruction, thereby reducing expiratory airflow(63). Ever-smoking was not independently associated with FVC or lung restriction after adjustment. Each additional year of age was associated with approximately 5% higher odds of COPD and approximately 2% higher odds of lung restriction. These findings are consistent with age-related reductions in lung elastic recoil, respiratory muscle strength and thoracic compliance(64–66).

This study has several strengths. We used harmonised spirometry protocols and standardised quality-control procedures across four African sites. The multisite design and rich demographic, behavioural, clinical and spirometric data enabled assessment of respiratory impairment alongside infectious and cardiometabolic comorbidities in underrepresented African populations. These data also provide a platform for longitudinal follow-up, development of African spirometric reference standards and genomic studies of respiratory disease and multimorbidity.

Several limitations should be considered. Data collection during the COVID-19 pandemic may have influenced recruitment. Pre-bronchodilator spirometry may have overestimated COPD prevalence by including reversible airflow obstruction, as post-bronchodilator testing was completed only in a small subset, while use of the fixed FEV_1_/FVC<0.70 threshold may have introduced age-related misclassification(41,42).

NHANES-III African American reference equations may not adequately represent variation in body size, body proportions, ancestry and early-life determinants of lung growth across African populations. In addition, race and ethnicity are social constructs rather than reliable biological proxies for lung function, and race-specific equations may mask reduced lung function arising from socioeconomic and environmental disadvantage(32,67). Exclusion of participants who did not meet repeatability thresholds, self-reported exposures, and residual between-site variability may also have influenced the estimates and their interpretation. The cross-sectional design precluded determination of temporality and limited causal interpretation of the observed associations.

Overall, our findings show that respiratory impairment is an important but under-characterised component of multimorbidity in African populations. The differing associations observed for COPD and lung restriction suggest that impaired lung function may reflect multiple infectious, cardiometabolic, ageing-related, environmental and social influences that vary across settings. African longitudinal studies using post-bronchodilator spirometry, repeated lung function assessment, improved measurement of tobacco, household air pollution, occupational, infectious, nutritional and cardiometabolic exposures, and locally appropriate reference standards are needed to clarify respiratory disease trajectories and guide interventions.

### Patient and public involvement statement

Community engagement was embedded in the broader AWI-Gen study. Before recruitment, AWI-Gen teams engaged community leaders and local stakeholders, including chiefs and elders, through community-entry meetings to explain the study and seek permission to approach community members. Community durbars, household and compound visits, group information sessions and individual informed consent were then used to support community understanding and participation. Feedback was implemented through community durbars and participant feedback activities, where aggregate study findings and health-education messages were shared with chiefs, elders, community members and study participants. Participants and community members were not directly involved in the design of the present spirometry analysis, statistical analysis, interpretation of results, or writing of this manuscript.

## Supporting information

Supplementary Tables S1-S4(participants characteristic and univariable analyses in AWI-Gen)

Supplementary Tables S5-S9(multivariable_linear_regression_for_FEV1_FVC_FEV1FVC_COPD_Lung_Restriction)

## Data Availability

All data produced in the present study are available upon reasonable request to the authors

## Contributors

Emmanuel Adonyo conducted the primary data analysis and drafted the manuscript. Anna L. Guyatt, Jing Chen, Martin Tobin and Lisa Micklesfield supervised the analysis, contributed to study design and interpretation of the findings, and critically revised the manuscript. Lisa K. Micklesfield, Michele Ramsay, Martin D. Tobin, Richard Packer, Palwende Romuald Boua, Anna L. Guyatt, and Godfred Agongo developed the spirometry standard operating procedures, and Lisa K. Micklesfield also coordinated data collection across AWI-Gen sites. Catherine John and Richard Packer contributed to spirometry implementation and quality assurance. Michele Ramsay oversaw the broader AWI-Gen study and coordinated data sharing. Lisa K. Micklesfield and Martin D. Tobin secured funding, provided strategic oversight, and critically reviewed the manuscript. All authors contributed to interpretation of the data and approved the final manuscript.

## Funding

This work was partially supported by the National Institute for Health Research (NIHR) Leicester Biomedical Research Centre (NIHR203327), and Martin Tobin is funded by an NIHR Senior Investigator Award. Data collection was partly supported by the AWI-Gen study (NHGRI; grant number U54HG006938). The views expressed are those of the author(s) and not necessarily those of the National Health Service, the NIHR, the Department of Health and Social Care in the UK, or the NIH in the USA.

This study was supported by a Wellcome Trust Discovery Award (WT225221/Z/22/Z).

Emmanuel Adonyo was funded by a Wellcome Trust PhD studentship as part of the Wellcome Trust Genetic Epidemiology and Public Health Genomics Doctoral Training Programme (218505/Z/19/Z).

This work was supported by the Academy of Medical Sciences and the UK Government Department for Science, Innovation and Technology through a GCRF Networking Grant [GCRFNGR3\1333].

Lisa K. Micklesfield is supported by the South African Medical Research Council.

## Notes

### Competing Interest Statement

The authors have declared no competing interest.

