## Supplementary Tables S1-S4(participants characteristic and univariable analyses in AWI-Gen) for "Lung function variability and the burden of COPD and lung restriction in South Africa and Burkina Faso: results from the AWI-Gen study"

**Supplementary information**

a) **Table S1** showing n (%), Mean (SD), and Median (Q1, Q3) the AWI-Gen population characteristics

| **Characteristic** | **Overall**  N = 1,619 | **Agincourt**  N = 130 | **DIMAMO**  N = 49 | **Nanoro**  N = 575 | **Soweto**  N = 865 |
| --- | --- | --- | --- | --- | --- |
| Date | 2019-01-24 to  2022-04-13 | 2019-11-18 to  2020-02-19 | 2019-04-25 to  2020-03-17 | 2020-12-22 to  2022-04-13 | 2019-01-24 to  2020-03-18 |
| Sex |  |  |  |  |  |
| Female | 936 (57.8%) | 86 (66.2%) | 30 (61.2%) | 338 (58.8%) | 482 (55.7%) |
| Male | 683 (42.2%) | 44 (33.8%) | 19 (38.8%) | 237 (41.2%) | 383 (44.3%) |
| Smoking Status |  |  |  |  |  |
| Ever-smoked | 352 (21.7%) | 8 (6.2%) | 10 (20.4%) | 53 (9.2%) | 281 (32.5%) |
| Never-smoked | 1,267 (78.3%) | 122 (93.8%) | 39 (79.6%) | 522 (90.8%) | 584 (67.5%) |
| Age (years) |  |  |  |  |  |
| Mean (SD) | 55.4 (6.3) | 60.8 (8.9) | 58.1 (6.5) | 54.5 (5.5) | 55.1 (5.9) |
| Median (Q1, Q3) | 55 (50.0, 60.0) | 62 (54.0, 68.0) | 59 (53.0, 62.0) | 54 (50.0, 59.0) | 55 (50.0, 60.0) |
| Height (m) |  |  |  |  |  |
| Min to Max | 1.4 to 1.9 | 1.5 to 1.9 | 1.5 to 1.8 | 1.5 to 1.9 | 1.4 to 1.9 |
| Mean (SD) | 1.6 (0.1) | 1.6 (0.1) | 1.6 (0.1) | 1.7 (0.1) | 1.6 (0.1) |
| Median (Q1, Q3) | 1.6 (1.6, 1.7) | 1.6 (1.6, 1.7) | 1.6 (1.6, 1.7) | 1.7 (1.6, 1.7) | 1.6 (1.6, 1.7) |
| Weight (kg) |  |  |  |  |  |
| Mean (SD) | 72.1 (19.7) | 76.4 (18.8) | 72.6 (18.9) | 58.6 (11.1) | 80.4 (19.4) |
| Median (Q1, Q3) | 69.1 (56.8, 84.1) | 75.1 (63.0, 87.0) | 68.0 (58.7, 86.1) | 56.8 (50.4, 65.0) | 78.7 (66.4, 92.7) |
| Best FEV1 (L) |  |  |  |  |  |
| Mean (SD) | 2.2 (0.6) | 2.0 (0.5) | 2.0 (0.7) | 2.0 (0.6) | 2.3 (0.6) |
| Median (Q1, Q3) | 2.1 (1.7, 2.6) | 2.0 (1.6, 2.3) | 2.0 (1.6, 2.4) | 1.9 (1.5, 2.3) | 2.3 (1.9, 2.8) |
| Best FVC (L) |  |  |  |  |  |
| Mean (SD) | 2.8 (0.8) | 2.6 (0.7) | 2.7 (0.8) | 2.6 (0.7) | 3.0 (0.8) |
| Median (Q1, Q3) | 2.7 (2.3, 3.3) | 2.5 (2.1, 3.0) | 2.6 (2.1, 3.3) | 2.5 (2.1, 3.1) | 2.9 (2.4, 3.5) |
| FEV1/FVC |  |  |  |  |  |
| Mean (SD) | 0.8 (0.1) | 0.8 (0.1) | 0.7 (0.1) | 0.8 (0.1) | 0.8 (0.1) |
| Median (Q1, Q3) | 0.8 (0.7, 0.8) | 0.8 (0.7, 0.8) | 0.8 (0.7, 0.8) | 0.8 (0.7, 0.8) | 0.8 (0.8, 0.8) |
| Best PEF (L/m) |  |  |  |  |  |
| Mean (SD) | 341.9 (129.5) | 317.3 (107.2) | 295.6 (114.4) | 275.3 (108.2) | 392.5 (124.3) |
| Median (Q1, Q3) | 332.3 (247.2, 425.3) | 316.1 (236.8, 387.4) | 288.3 (200.7, 368.4) | 264.4 (194.0, 344.6) | 385.6 (302.9, 470.6) |
| Predicted FEV1 |  |  |  |  |  |
| Mean (SD) | 2.5 (0.5) | 2.3 (0.5) | 2.3 (0.4) | 2.5 (0.5) | 2.4 (0.5) |
| Median (Q1, Q3) | 2.4 (2.0, 2.9) | 2.2 (1.9, 2.5) | 2.3 (1.9, 2.6) | 2.4 (2.1, 2.9) | 2.4 (2.0, 2.9) |
| Predicted FVC |  |  |  |  |  |
| Mean (SD) | 3.1 (0.7) | 2.9 (0.7) | 2.9 (0.6) | 3.2 (0.6) | 3.1 (0.7) |
| Median (Q1, Q3) | 3.0 (2.6, 3.6) | 2.8 (2.4, 3.3) | 2.9 (2.4, 3.4) | 3.0 (2.7, 3.7) | 3.0 (2.5, 3.6) |
| Predicted FEV1/FVC |  |  |  |  |  |
| Mean (SD) | 0.8 (0.0) | 0.8 (0.0) | 0.8 (0.0) | 0.8 (0.0) | 0.8 (0.0) |
| Median (Q1, Q3) | 0.8 (0.8, 0.8) | 0.8 (0.8, 0.8) | 0.8 (0.8, 0.8) | 0.8 (0.8, 0.8) | 0.8 (0.8, 0.8) |
| Predicted PEF |  |  |  |  |  |
| Mean (SD) | 6.7 (1.3) | 6.2 (1.4) | 6.3 (1.2) | 6.8 (1.3) | 6.7 (1.3) |
| Median (Q1, Q3) | 6.3 (5.6, 7.8) | 6.0 (5.2, 7.2) | 6.1 (5.2, 7.3) | 6.4 (5.8, 7.9) | 6.3 (5.5, 7.8) |
| % Predicted FEV1 |  |  |  |  |  |
| Mean (SD) | 88.7 (20.6) | 88.1 (18.0) | 89.3 (25.2) | 77.0 (17.0) | 96.6 (19.1) |
| Median (Q1, Q3) | 89.6 (75.6, 102.4) | 89.1 (74.7, 100.0) | 91.4 (71.7, 108.8) | 78.5 (66.5, 89.3) | 97.4 (85.8, 108.7) |
| % Predicted FVC |  |  |  |  |  |
| Mean (SD) | 90.8 (18.7) | 90.3 (17.3) | 93.8 (22.0) | 81.2 (16.6) | 97.0 (17.3) |
| Median (Q1, Q3) | 91.1 (78.9, 102.7) | 90.4 (77.7, 101.0) | 97.1 (78.1, 109.7) | 82.1 (71.6, 92.7) | 97.3 (86.1, 108.2) |
| % Predicted FEV1/FVC |  |  |  |  |  |
| Mean (SD) | 97.0 (12.0) | 96.9 (9.4) | 93.9 (12.9) | 94.9 (15.4) | 98.6 (9.1) |
| Median (Q1, Q3) | 98.8 (92.2, 103.9) | 98.2 (93.9, 102.0) | 96.2 (89.7, 101.2) | 96.3 (87.8, 103.2) | 100.3 (94.8, 104.5) |
| % Predicted PEF |  |  |  |  |  |
| Mean (SD) | 5,150.2 (1,747.4) | 5,106.7 (1,448.1) | 4,784.2 (1,723.0) | 4,002.8 (1,276.3) | 5,940.3 (1,627.5) |
| Median (Q1, Q3) | 5,071.4 (3,846.5, 6,321.0) | 5,107.1 (3,986.4, 5,987.6) | 4,852.6 (3,504.9, 6,040.2) | 3,932.0 (3,012.5, 4,885.0) | 5,932.5 (4,845.6, 7,082.9) |
| LLN FEV1 |  |  |  |  |  |
| Mean (SD) | 1.8 (0.4) | 1.6 (0.4) | 1.6 (0.4) | 1.9 (0.4) | 1.8 (0.4) |
| Median (Q1, Q3) | 1.7 (1.5, 2.1) | 1.6 (1.3, 1.8) | 1.7 (1.3, 1.9) | 1.8 (1.5, 2.2) | 1.7 (1.4, 2.1) |
| LLN FVC |  |  |  |  |  |
| Mean (SD) | 2.3 (0.5) | 2.2 (0.6) | 2.2 (0.5) | 2.4 (0.5) | 2.3 (0.6) |
| Median (Q1, Q3) | 2.2 (1.9, 2.8) | 2.1 (1.7, 2.5) | 2.2 (1.7, 2.5) | 2.3 (2.0, 2.8) | 2.2 (1.9, 2.8) |
| LLN FEV1/FVC |  |  |  |  |  |
| Mean (SD) | 0.7 (0.0) | 0.7 (0.0) | 0.7 (0.0) | 0.7 (0.0) | 0.7 (0.0) |
| Median (Q1, Q3) | 0.7 (0.7, 0.7) | 0.7 (0.7, 0.7) | 0.7 (0.7, 0.7) | 0.7 (0.7, 0.7) | 0.7 (0.7, 0.7) |
| LLN PEF |  |  |  |  |  |
| Mean (SD) | 4.5 (1.0) | 4.1 (1.1) | 4.2 (0.9) | 4.6 (1.0) | 4.5 (1.0) |
| Median (Q1, Q3) | 4.3 (3.7, 5.4) | 4.0 (3.3, 4.8) | 4.1 (3.3, 5.0) | 4.3 (3.8, 5.5) | 4.2 (3.6, 5.4) |
| Quality Grade |  |  |  |  |  |
| A | 692 (42.7%) | 57 (43.8%) | 17 (34.7%) | 156 (27.1%) | 462 (53.4%) |
| B | 316 (19.5%) | 29 (22.3%) | 7 (14.3%) | 109 (19.0%) | 171 (19.8%) |
| C | 139 (8.6%) | 14 (10.8%) | 6 (12.2%) | 62 (10.8%) | 57 (6.6%) |
| C2 | 56 (3.5%) | 9 (6.9%) | 0 (0.0%) | 22 (3.8%) | 25 (2.9%) |
| D1 | 262 (16.2%) | 15 (11.5%) | 7 (14.3%) | 144 (25.0%) | 96 (11.1%) |
| D2 | 154 (9.5%) | 6 (4.6%) | 12 (24.5%) | 82 (14.3%) | 54 (6.2%) |
| GOLD 1+ |  |  |  |  |  |
| 0 | 1,358 (83.9%) | 115 (88.5%) | 37 (75.5%) | 434 (75.5%) | 772 (89.2%) |
| 1 | 261 (16.1%) | 15 (11.5%) | 12 (24.5%) | 141 (24.5%) | 93 (10.8%) |
| GOLD 2+ |  |  |  |  |  |
| 0 | 1,425 (88.0%) | 119 (91.5%) | 41 (83.7%) | 458 (79.7%) | 807 (93.3%) |
| 1 | 194 (12.0%) | 11 (8.5%) | 8 (16.3%) | 117 (20.3%) | 58 (6.7%) |
| Restriction |  |  |  |  |  |
| 0 | 1,274 (78.7%) | 99 (76.2%) | 41 (83.7%) | 367 (63.8%) | 767 (88.7%) |
| 1 | 345 (21.3%) | 31 (23.8%) | 8 (16.3%) | 208 (36.2%) | 98 (11.3%) |

b) **Table S2:** Univariable linear regression for FEV_1_, FVC and FEV_1_/FVC in AWI-Gen. TB – tuberculosis, SES – Social economic quintiles, CKD – chronic kidney diseases.

|  |  | **FEV_1_ (L)** | | | **FVC (L)** | | | **FEV_1_/FVC** | | |
| --- | --- | --- | --- | --- | --- | --- | --- | --- | --- | --- |
|  | N | Beta (L) | 95% CI | p-value | Beta (L) | 95% CI | p-value | Beta | 95% CI | p-value |
| Age (years) | 1,619 | -0.025 | -0.030, -0.02 | 7.40x10^-24^ | -0.026 | -0.032, -0.02 | 9.70x10^-17^ | -0.002 | -0.003, -0.001 | 2.48x10^-6^ |
| Height (metres) | 1,619 | 3.95 | 3.65, 4.26 | 9.87x10^-121^ | 5.52 | 5.17, 5.88 | 7.20x10^-164^ | -0.115 | -0.169, -0.062 | 2.71x10^-5^ |
| Sex (Ref=female) | 936/683 | 0.737 | 0.685, 0.790 | 2.51x10^-137^ | 1.02 | 0.960, 1.08 | 9.01x10^-184^ | -0.020 | -0.030, -0.011 | 3.09x10^-5^ |
| Smoking status  (Ref=never-smokers) | 1,267/352 | 0.560 | 0.489, 0.631 | 2.51x10^-50^ | 0.803 | 0.718, 0.889 | 2.22x10^-69^ | -0.023 | -0.035, -0.012 | 6.74x10^-5^ |
| SES quintile | 1,619 | 0.112 | 0.087, 0.137 | 7.36x10^-18^ | 0.133 | 0.102, 0.165 | 8.69x10^-17^ | 0.002 | -0.002, 0.006 | 0.265 |
| Study site | 1,619 |  | | | | | | | | |
| (Ref=Soweto) | 866 |  | | | | | | | | |
| Agincourt | 130 | -0.348 | -0.462, -0.234 | 2.65x10^-9^ | -0.367 | -0.510, -0.224 | 5.04x10^-7^ | -0.023 | -0.040, -0.005 | 0.0117 |
| Dimamo | 49 | -0.339 | -0.517, -0.162 | 1.82x10^-4^ | -0.279 | -0.501, -0.057 | 0.0138 | -0.042 | -0.070, -0.015 | 2.52x10^-3^ |
| Nanoro | 575 | -0.392 | -0.457, -0.327 | 5.20x10^-31^ | -0.392 | -0.473, -0.311 | 1.08x10^-20^ | -0.028 | -0.038, -0.018 | 6.31x10^-8^ |
| Solid fuel exposure | 1,540 |  | | | | | | | | |
| (Ref=“No solid fuel use”) | 946 |  | | | | | | | | |
| Solid fuel use with  kitchen ventilation | 553 | -0.382 | -0.448, -0.317 | 2.57x10^-29^ | -0.391 | -0.473, -0.309 | 1.89x10^-20^ | -0.025 | -0.035, -0.015 | 1.59x10^-6^ |
| Solid fuel use with  no kitchen ventilation | 41 | -0.288 | -0.483, -0.094 | 3.72x10^-3^ | -0.218 | -0.461, 0.025 | 0.0784 | -0.039 | -0.070, -0.009 | 0.0109 |
| TB (Ref=no) | 1,499/118 | 0.145 | 0.025, 0.266 | 0.0182 | 0.273 | 0.125, 0.421 | 3.06x10^-4^ | -0.025 | -0.043, -0.006 | 7.92x10^-3^ |
| Diabetes (Ref=no) | 1,447/128 | -0.010 | -0.127, 0.107 | 0.865 | -0.113 | -0.257, 0.031 | 0.123 | 0.027 | 0.010, 0.045 | 1.96 x10^-3^ |
| Cardiovascular  disease (Ref=no) | 1,404/126 | -0.127 | -0.244, -0.011 | 0.0319 | -0.224 | -0.367, -0.081 | 2.21x10^-3^ | 0.020 | 0.002, 0.038 | 0.026 |
| Hypertension (Ref=no) | 1,083/507 | -0.063 | -0.131, 0.005 | 0.069 | -0.137 | -0.221, -0.053 | 1.35x10^-3^ | 0.014 | 0.004, 0.024 | 7.54 x10^-3^ |
| CKD(Ref=no) | 1,553/60 | 0.084 | -0.083, 0.250 | 0.324 | 0.067 | -0.138, 0.272 | 0.521 | 0.010 | -0.014, 0.035 | 0.414 |
| HIV status (Ref=no) | 899/190 | 0.061 | -0.039, 0.161 | 0.230 | 0.117 | -0.009, 0.242 | 0.0677 | -0.008 | -0.021, 0.005 | 0.218 |

c) **Table S3:** Univariable logistic regression for COPD in AWI-Gen

| Characteristic | N | OR | 95% CI | p-value |
| --- | --- | --- | --- | --- |
| Age (Years) | 1,619 | 1.0 | 1.0, 1.1 | 2.00e-03 |
| Height (per 10 cm increase) | 1,619 | 1.3 | 1.1, 1.5 | 1.35e-03 |
| Sex (Male) | 683 | 1.4 | 1.1, 1.9 | 9.94e-03 |
| Smoking status (ever-smokers) | 352 | 1.5 | 1.1, 2.0 | 1.28e-02 |
| SES Quintile | 1,619 | 0.90 | 0.70, 1.2 | 4.25e-01 |
| Study site (Soweto) | 865 | — | — | NA |
| Agincourt | 130 | 1.1 | 0.59, 1.9 | 7.88e-01 |
| DIMAMO | 49 | 2.7 | 1.3, 5.2 | 4.64e-03 |
| Nanoro | 575 | 2.7 | 2.0, 3.6 | 1.24e-11 |
| No solid fuel use | 945 | — | — | NA |
| Solid fuel use with kitchen ventilation | 553 | 2.5 | 1.9, 3.4 | 9.53e-11 |
| Solid fuel use with no kitchen ventilation | 41 | 2.9 | 1.4, 5.9 | 3.40e-03 |
| TB (cases) | 118 | 2.1 | 1.3, 3.1 | 9.48e-04 |
| Diabetes (cases) | 128 | 0.44 | 0.21, 0.81 | 1.40e-02 |
| CVD (cases) | 126 | 0.76 | 0.43, 1.3 | 3.27e-01 |
| Hypertension (cases) | 508 | 0.74 | 0.54, 0.99 | 4.72e-02 |
| CKD (cases) | 60 | 0.68 | 0.28, 1.4 | 3.42e-01 |
| HIV Status (cases) | 190 | 1.3 | 0.81, 2.0 | 2.62e-01 |
| Abbreviations: CI = Confidence Interval, OR = Odds Ratio | | | | |

d) **Table S4:** Univariable logistic regression for lung restriction in AWI-Gen

| Characteristic | N | OR | 95% CI | p-value |
| --- | --- | --- | --- | --- |
| Age (Years) | 1,619 | 1.0 | 1.0, 1.0 | 1.39e-01 |
| Height (per 10 cm increase) | 1,619 | 1.1 | 0.99, 1.3 | 7.73e-02 |
| Sex (Male) | 683 | 0.85 | 0.67, 1.1 | 1.95e-01 |
| Smoking status (ever-smokers) | 352 | 0.58 | 0.41, 0.79 | 7.94e-04 |
| SES Quintile | 1,619 | 0.75 | 0.59, 0.93 | 1.07e-02 |
| Study site (Soweto) | 865 | — | — | NA |
| Agincourt | 130 | 2.5 | 1.5, 3.8 | 1.12e-04 |
| DIMAMO | 49 | 1.5 | 0.65, 3.2 | 2.91e-01 |
| Nanoro | 575 | 4.4 | 3.4, 5.8 | 3.59e-27 |
| No solid fuel use | 945 | — | — | NA |
| Solid fuel use with kitchen ventilation | 553 | 4.1 | 3.2, 5.3 | 4.37e-26 |
| Solid fuel use with no kitchen ventilation | 41 | 3.8 | 1.9, 7.3 | 1.12e-04 |
| TB (cases) | 118 | 0.79 | 0.47, 1.3 | 3.39e-01 |
| Diabetes (cases) | 128 | 0.90 | 0.56, 1.4 | 6.54e-01 |
| CVD (cases) | 126 | 1.1 | 0.72, 1.7 | 5.71e-01 |
| Hypertension (cases) | 508 | 0.82 | 0.63, 1.1 | 1.49e-01 |
| CKD (cases) | 60 | 0.82 | 0.40, 1.5 | 5.65e-01 |
| HIV Status (cases) | 190 | 0.99 | 0.62, 1.5 | 9.60e-01 |
| Abbreviations: CI = Confidence Interval, OR = Odds Ratio | | | | |
