## Supplementary Tables S5-S9(multivariable_linear_regression_for_FEV1_FVC_FEV1FVC_COPD_Lung_Restriction) for "Lung function variability and the burden of COPD and lung restriction in South Africa and Burkina Faso: results from the AWI-Gen study"

Note: The base model includes age, height, sex, smoking status, study site, and socioeconomic status (SES, modelled as a continuous quintile score). Each subsequent column represents a separate model in which one additional predictor (solid fuel exposure, tuberculosis, diabetes, cardiovascular disease, hypertension, chronic kidney disease, or HIV status) is added individually to the base model. Coefficients are presented as β estimates with 95% confidence intervals. Sample size (N) and R² are reported for each model and may vary across columns due to differences in data completeness for the added predictor.

a) **Table S5** Multivariable linear regression analyses of FEV_1_ in AWI-Gen participants.

|  | Base model | Base model +  Solid fuel  exposure | Base model +  TB | Base model +  Diabetes | Base model +  CVD | Base model +  Hypertension | Base model +  CKD | Base model +  HIV Status |
| --- | --- | --- | --- | --- | --- | --- | --- | --- |
|  | Beta  95% [CI] | Beta  95% [CI] | Beta  95% [CI] | Beta  95% [CI] | Beta  95% [CI] | Beta  95% [CI] | Beta  95% [CI] | Beta  95% [CI] |
| Age  (Years) | **-0.02**  **[-0.03, -0.02]** | **-0.02**  **[-0.03, -0.02]** | **-0.02**  **[-0.03, -0.02]** | **-0.02**  **[-0.03, -0.02]** | **-0.03**  **[-0.03, -0.02]** | **-0.02**  **[-0.03, -0.02]** | **-0.02**  **[-0.03, -0.02]** | **-0.03**  **[-0.03, -0.02]** |
| Height  (metres) | **2.54**  **[2.20, 2.89]** | **2.57**  **[2.21, 2.93]** | **2.55**  **[2.21, 2.90]** | **2.51**  **[2.16, 2.86]** | **2.58**  **[2.23, 2.94]** | **2.53**  **[2.18, 2.88]** | **2.55**  **[2.21, 2.90]** | **2.39**  **[1.95, 2.82]** |
| Sex  (Male) | **0.45**  **[0.38, 0.51]** | **0.46**  **[0.39, 0.53]** | **0.44**  **[0.38, 0.51]** | **0.45**  **[0.38, 0.52]** | **0.45**  **[0.38, 0.52]** | **0.45**  **[0.38, 0.52]** | **0.44**  **[0.38, 0.51]** | **0.46**  **[0.37, 0.55]** |
| Smoking status  (ever-smokers) | **-0.08**  **[-0.15, -0.02]** | **-0.09**  **[-0.16, -0.02]** | **-0.06**  **[-0.13, 0.00]** | **-0.08**  **[-0.15, -0.01]** | **-0.09**  **[-0.16, -0.02]** | **-0.09**  **[-0.16, -0.03]** | **-0.08**  **[-0.15, -0.02]** | **-0.08**  **[-0.16, -0.00]** |
| Study site (Soweto) |  |  |  |  |  |  |  |  |
| Agincourt | **-0.15**  **[-0.24, -0.05]** | -0.08  [-0.22, 0.06] | **-0.15**  **[-0.25, -0.06]** | **-0.15**  **[-0.25, -0.06]** | **-0.13**  **[-0.23, -0.02]** | **-0.15**  **[-0.25, -0.06]** | **-0.15**  **[-0.24, -0.05]** | -0.11  [-0.22, 0.01] |
| DIMAMO | **-0.18**  **[-0.31, -0.05]** | **-0.16**  **[-0.31, -0.01]** | **-0.19**  **[-0.31, -0.06]** | **-0.17**  **[-0.30, -0.04]** | **-0.20**  **[-0.33, -0.06]** | **-0.21**  **[-0.34, -0.07]** | **-0.19**  **[-0.33, -0.06]** | **-0.19**  **[-0.33, -0.05]** |
| Nanoro | **-0.46**  **[-0.51, -0.41]** | **-0.40**  **[-0.59, -0.22]** | **-0.48**  **[-0.53, -0.43]** | **-0.46**  **[-0.51, -0.41]** | **-0.47**  **[-0.52, -0.42]** | **-0.50**  **[-0.55, -0.44]** | **-0.46**  **[-0.52, -0.41]** | **-0.44**  **[-0.54, -0.33]** |
| SES  Quintile | **0.06**  **[0.01, 0.11]** | **0.06**  **[0.01, 0.10]** | **0.06**  **[0.02, 0.11]** | **0.06**  **[0.01, 0.10]** | **0.06**  **[0.01, 0.10]** | **0.06**  **[0.01, 0.11]** | **0.06**  **[0.01, 0.11]** | 0.06  [-0.00, 0.11] |
| Solid fuel exposure |  |  |  |  |  |  |  |  |
| Solid fuel use with  kitchen ventilation |  | -0.06  [-0.25, 0.12] |  |  |  |  |  |  |
| Solid fuel use with  no kitchen ventilation |  | -0.04  [-0.27, 0.18] |  |  |  |  |  |  |
| TB  (cases) |  |  | **-0.18**  **[-0.27, -0.10]** |  |  |  |  |  |
| Diabetes  (cases) |  |  |  | -0.01  [-0.09, 0.07] |  |  |  |  |
| CVD  (cases) |  |  |  |  | -0.03  [-0.11, 0.05] |  |  |  |
| Hypertension  (cases) |  |  |  |  |  | **-0.09**  **[-0.14, -0.04]** |  |  |
| CKD  (cases) |  |  |  |  |  |  | -0.03  [-0.14, 0.09] |  |
| HIV Status  (cases) |  |  |  |  |  |  |  | -0.05  [-0.13, 0.02] |
| Overall N | 1619 | 1540 | 1617 | 1575 | 1530 | 1590 | 1613 | 1089 |
| R^2^ | 0.520 | 0.523 | 0.524 | 0.519 | 0.521 | 0.525 | 0.520 | 0.477 |

b) **Table S6** Multivariable linear regression analyses of FVC in AWI-Gen participants.

|  | Base model | Base model +  Solid fuel  exposure | Base model +  TB | Base model +  Diabetes | Base model +  CVD | Base model +  Hypertension | Base model +  CKD | Base model +  HIV Status |
| --- | --- | --- | --- | --- | --- | --- | --- | --- |
|  | Beta  95% [CI] | Beta  95% [CI] | Beta  95% [CI] | Beta  95% [CI] | Beta  95% [CI] | Beta  95% [CI] | Beta  95% [CI] | Beta  95% [CI] |
| Age (Years) | **-0.02**  **[-0.03, -0.02]** | **-0.02**  **[-0.03, -0.02]** | **-0.02**  **[-0.03, -0.02]** | **-0.02**  **[-0.03, -0.02]** | **-0.02**  **[-0.03, -0.02]** | **-0.02**  **[-0.03, -0.02]** | **-0.02**  **[-0.03, -0.02]** | **-0.03**  **[-0.03, -0.02]** |
| Height (metres) | **3.46**  **[3.06, 3.87]** | **3.52**  **[3.11, 3.94]** | **3.47**  **[3.07, 3.88]** | **3.45**  **[3.04, 3.86]** | **3.50**  **[3.09, 3.92]** | **3.45**  **[3.05, 3.86]** | **3.47**  **[3.07, 3.88]** | **3.30**  **[2.81, 3.80]** |
| Sex (Male) | **0.58**  **[0.50, 0.66]** | **0.58**  **[0.50, 0.66]** | **0.58**  **[0.50, 0.65]** | **0.58**  **[0.50, 0.66]** | **0.57**  **[0.49, 0.65]** | **0.58**  **[0.50, 0.66]** | **0.58**  **[0.50, 0.65]** | **0.63**  **[0.52, 0.73]** |
| Smoking status  (ever-smokers) | 0.01  [-0.07, 0.09] | 0.01  [-0.07, 0.09] | 0.03  [-0.05, 0.10] | 0.01  [-0.07, 0.09] | 0.02  [-0.06, 0.09] | -0.01  [-0.08, 0.07] | 0.01  [-0.07, 0.09] | 0.01  [-0.08, 0.10] |
| Study site (Soweto) |  | | | | | | | |
| Agincourt | **-0.13**  **[-0.24, -0.02]** | -0.09  [-0.26, 0.07] | **-0.14**  **[-0.25, -0.03]** | **-0.15**  **[-0.26, -0.04]** | -0.10  [-0.23, 0.02] | **-0.14**  **[-0.25, -0.03]** | **-0.14**  **[-0.25, -0.03]** | -0.09  [-0.23, 0.04] |
| DIMAMO | -0.09  [-0.24, 0.06] | -0.06  [-0.23, 0.12] | -0.09  [-0.24, 0.06] | -0.07  [-0.22, 0.08] | -0.10  [-0.25, 0.06] | -0.12  [-0.27, 0.03] | -0.10  [-0.25, 0.06] | -0.09  [-0.24, 0.07] |
| Nanoro | **-0.46**  **[-0.52, -0.40]** | **-0.38**  **[-0.60, -0.16]** | **-0.47**  **[-0.53, -0.41]** | **-0.47**  **[-0.53, -0.41]** | **-0.46**  **[-0.52, -0.40]** | **-0.52**  **[-0.58, -0.46]** | **-0.46**  **[-0.52, -0.40]** | **-0.46**  **[-0.58, -0.34]** |
| SES  Quintile | **0.06**  **[0.01, 0.12]** | **0.06**  **[0.01, 0.12]** | **0.06**  **[0.01, 0.12]** | **0.06**  **[0.01, 0.12]** | **0.06**  **[0.01, 0.12]** | **0.07**  **[0.01, 0.12]** | **0.06**  **[0.01, 0.12]** | 0.05  [-0.01, 0.12] |
| Solid fuel exposure |  | | | | | | | |
| Solid fuel use with  kitchen ventilation |  | -0.09  [-0.30, 0.12] |  |  |  |  |  |  |
| Solid fuel use with  no kitchen ventilation |  | -0.01  [-0.27, 0.25] |  |  |  |  |  |  |
| TB  (cases) |  |  | **-0.13**  **[-0.23, -0.03]** |  |  |  |  |  |
| Diabetes  (cases) |  |  |  | **-0.11**  **[-0.20, -0.01]** |  |  |  |  |
| CVD  (cases) |  |  |  |  | -0.09  [-0.18, 0.00] |  |  |  |
| Hypertension  (cases) |  |  |  |  |  | **-0.15**  **[-0.21, -0.09]** |  |  |
| CKD  (cases) |  |  |  |  |  |  | -0.05  [-0.18, 0.09] |  |
| HIV Status  (cases) |  |  |  |  |  |  |  | -0.00  [-0.09, 0.08] |
| Overall N | 1619 | 1540 | 1617 | 1575 | 1530 | 1590 | 1613 | 1089 |
| R^2^ | 0.569 | 0.572 | 0.570 | 0.574 | 0.570 | 0.579 | 0.568 | 0.562 |

c) **Table S7** Multivariable linear regression analyses of FEV1/FVC in AWI-Gen participants.

|  | Base model | Base model +  Solid fuel  exposure | Base model +  TB | Base model +  Diabetes | Base model +  CVD | Base model +  Hypertension | Base model +  CKD | Base model +  HIV Status |
| --- | --- | --- | --- | --- | --- | --- | --- | --- |
|  | Beta  95% [CI] | Beta  95% [CI] | Beta  95% [CI] | Beta  95% [CI] | Beta  95% [CI] | Beta  95% [CI] | Beta  95% [CI] | Beta  95% [CI] |
| Age (Years) | **-0.00**  **[-0.00, -0.00]** | **-0.00**  **[-0.00, -0.00]** | **-0.00**  **[-0.00, -0.00]** | **-0.00**  **[-0.00, -0.00]** | **-0.00**  **[-0.00, -0.00]** | **-0.00**  **[-0.00, -0.00]** | **-0.00**  **[-0.00, -0.00]** | **-0.00**  **[-0.00, -0.00]** |
| Height (metres) | -0.04  [-0.12, 0.03] | -0.05  [-0.13, 0.02] | -0.04  [-0.11, 0.03] | -0.05  [-0.13, 0.02] | -0.04  [-0.12, 0.03] | -0.05  [-0.12, 0.03] | -0.04  [-0.11, 0.03] | -0.06  [-0.14, 0.01] |
| Sex (Male) | -0.00  [-0.02, 0.01] | -0.00  [-0.01, 0.01] | -0.00  [-0.02, 0.01] | -0.00  [-0.02, 0.01] | -0.00  [-0.02, 0.01] | -0.00  [-0.02, 0.01] | -0.00  [-0.02, 0.01] | -0.01  [-0.03, 0.00] |
| Smoking status  (ever-smokers) | **-0.03**  **[-0.05, -0.02]** | **-0.03**  **[-0.05, -0.02]** | **-0.03**  **[-0.04, -0.01]** | **-0.03**  **[-0.05, -0.02]** | **-0.04**  **[-0.05, -0.02]** | **-0.03**  **[-0.04, -0.02]** | **-0.03**  **[-0.05, -0.02]** | **-0.03**  **[-0.04, -0.01]** |
| Study site (Soweto) |  | | | | | | | |
| Agincourt | -0.02  [-0.04, 0.00] | -0.00  [-0.03, 0.03] | -0.02  [-0.04, 0.00] | -0.02  [-0.04, 0.00] | -0.02  [-0.04, 0.00] | -0.02  [-0.04, 0.00] | -0.02  [-0.04, 0.00] | -0.01  [-0.03, 0.01] |
| DIMAMO | **-0.04**  **[-0.07, -0.01]** | **-0.04**  **[-0.07, -0.01]** | **-0.04**  **[-0.07, -0.01]** | **-0.04**  **[-0.07, -0.01]** | **-0.04**  **[-0.07, -0.01]** | **-0.04**  **[-0.07, -0.01]** | **-0.04**  **[-0.07, -0.01]** | **-0.04**  **[-0.07, -0.02]** |
| Nanoro | **-0.04**  **[-0.05, -0.02]** | -0.04  [-0.08, 0.00] | **-0.04**  **[-0.05, -0.03]** | **-0.03**  **[-0.04, -0.02]** | **-0.04**  **[-0.05, -0.03]** | **-0.03**  **[-0.04, -0.02]** | **-0.04**  **[-0.05, -0.02]** | -0.02  [-0.03, 0.00] |
| SES  Quintile | 0.00  [-0.01, 0.01] | 0.00  [-0.01, 0.01] | 0.00  [-0.01, 0.01] | 0.00  [-0.01, 0.01] | 0.00  [-0.01, 0.01] | 0.00  [-0.01, 0.01] | 0.00  [-0.01, 0.01] | 0.01  [-0.00, 0.02] |
| Solid fuel exposure |  | | | | | | | |
| Solid fuel use with  kitchen ventilation |  | 0.00  [-0.03, 0.04] |  |  |  |  |  |  |
| Solid fuel use with  no kitchen ventilation |  | -0.01  [-0.06, 0.04] |  |  |  |  |  |  |
| TB  (cases) |  |  | **-0.03**  **[-0.05, -0.01]** |  |  |  |  |  |
| Diabetes  (cases) |  |  |  | 0.03  [0.01, 0.04] |  |  |  |  |
| CVD  (cases) |  |  |  |  | 0.02  [0.00, 0.03] |  |  |  |
| Hypertension  (cases) |  |  |  |  |  | 0.01  [-0.00, 0.02] |  |  |
| CKD  (cases) |  |  |  |  |  |  | 0.00  [-0.02, 0.03] |  |
| HIV Status  (cases) |  |  |  |  |  |  |  | **-0.01**  **[-0.03, -0.00]** |
| Overall N | 1619 | 1540 | 1617 | 1575 | 1530 | 1590 | 1613 | 1089 |
| R^2^ | 0.060 | 0.059 | 0.066 | 0.063 | 0.067 | 0.059 | 0.060 | 0.099 |

d) **Table S8** Multivariable logistic regression analyses of COPD in the AWI-Gen analytical population.

|  | Base model | Base model  + Solid  Fuel exposure | Base model  +  TB | Base model  +  Diabetes | Base model  +  CVD | Base model  +  Hyperte  nsion | Base model  +  CKD | Base model  +  HIV infection |
| --- | --- | --- | --- | --- | --- | --- | --- | --- |
|  | OR (95% CI) | OR (95% CI) | OR (95% CI) | OR (95% CI) | OR (95% CI) | OR (95% CI) | OR (95% CI) | OR (95% CI) |
| Age (years) | **1.05**  **[1.03, 1.07]** | **1.05**  **[1.02, 1.07]** | **1.05**  **[1.03, 1.08]** | **1.05**  **[1.03, 1.08]** | **1.06**  **[1.03, 1.09]** | **1.05**  **[1.02, 1.07]** | **1.05**  **[1.03, 1.07]** | **1.06**  **[1.03, 1.10]** |
| Height  (per 10 cm increase) | 1.12  [0.91, 1.39] | 1.14  [0.91, 1.42] | 1.11  [0.89, 1.38] | 1.19  [0.95, 1.48] | 1.13  [0.90, 1.42] | 1.14  [0.92, 1.42] | 1.11  [0.90, 1.38] | 1.15  [0.86, 1.54] |
| Sex (Male) | 0.93  [0.61, 1.41] | 0.85  [0.55, 1.30] | 0.94  [0.62, 1.43] | 0.87  [0.57, 1.33] | 0.89  [0.58, 1.38] | 0.89  [0.58, 1.36] | 0.94  [0.62, 1.43] | 1.33  [0.73, 2.44] |
| Ever smoking  (vs never smoking) | **2.21**  **[1.48, 3.32]** | **2.33**  **[1.55, 3.54]** | **1.96**  **[1.30, 2.97]** | **2.24**  **[1.48, 3.38]** | **2.50**  **[1.64, 3.81]** | **2.29**  **[1.52, 3.46]** | **2.22**  **[1.48, 3.33]** | **2.50**  **[1.52, 4.16]** |
| Study site (Soweto) |  | | | | | | | |
| Agincourt | 0.88  [0.44, 1.68] | 0.43  [0.10, 1.29] | 0.92  [0.46, 1.76] | 0.81  [0.39, 1.57] | 0.81  [0.32, 1.80] | 0.90  [0.45, 1.70] | 0.89  [0.45, 1.69] | 0.80  [0.34, 1.75] |
| DIMAMO | **2.58**  **[1.22, 5.13]** | **2.80**  **[1.19, 6.14]** | **2.63**  **[1.23, 5.30]** | **2.47**  **[1.13, 5.01]** | **2.38**  **[1.06, 4.96]** | **2.63**  **[1.24, 5.26]** | **2.67**  **[1.26, 5.34]** | **2.79**  **[1.28, 5.74]** |
| Nanoro | **3.38**  **[2.45, 4.69]** | **4.10**  **[1.37, 12.42]** | **3.82**  **[2.74, 5.36]** | **3.03**  **[2.18, 4.24]** | **3.65**  **[2.62, 5.13]** | **3.36**  **[2.37, 4.79]** | **3.37**  **[2.44, 4.70]** | **2.74**  **[1.46, 5.02]** |
| SES quintile  (continuous) | 0.86  [0.64, 1.16] | 0.87  [0.65, 1.17] | 0.85  [0.63, 1.14] | 0.90  [0.67, 1.22] | 0.88  [0.65, 1.19] | 0.86  [0.64, 1.16] | 0.87  [0.65, 1.16] | 0.81  [0.55, 1.20] |
| Solid fuel exposure  (reference:  no solid fuel use) |  | | | | | | | |
| Solid fuel use with  kitchen ventilation |  | 0.82  [0.28, 2.38] |  |  |  |  |  |  |
| Solid fuel use with  no kitchen ventilation |  | 0.97  [0.27, 3.36] |  |  |  |  |  |  |
| Prior TB (yes vs no) |  |  | **2.86**  **[1.75, 4.59]** |  |  |  |  |  |
| Diabetes (yes vs no) |  |  |  | **0.50**  **[0.24, 0.94**] |  |  |  |  |
| CVD (yes vs no) |  |  |  |  | 0.80  [0.44, 1.37] |  |  |  |
| Hypertension  (yes vs no) |  |  |  |  |  | 1.05  [0.74, 1.48] |  |  |
| CKD (yes vs no) |  |  |  |  |  |  | 0.93  [0.37, 2.01] |  |
| HIV infection  (yes vs no) |  |  |  |  |  |  |  | **1.67**  **[1.01, 2.72]** |
| Overall N | 1619 | 1539 | 1617 | 1575 | 1530 | 1590 | 1613 | 1089 |
| *Note: Values are adjusted ORs (95% CIs). Base models included age, height, sex, ever smoking, study site and SES (continuous quintile score); each additional exposure was modelled separately, so N varies with data completeness. Bold denotes associations for which the 95% CI excluded the null value before rounding; some confidence limits round to 1.00. TB, tuberculosis; CVD, cardiovascular disease; CKD, chronic kidney disease; HIV, human immunodeficiency virus.* | | | | | | | | |

e) **Table S9** Multivariable logistic regression analyses of lung restriction in the AWI-Gen analytical population.

|  | Base model | Base model  + Solid  Fuel exposure | Base model  +  TB | Base model  +  Diabetes | Base model  +  CVD | Base model  +  Hyperte  nsion | Base model  +  CKD | Base model  +  HIV infection |
| --- | --- | --- | --- | --- | --- | --- | --- | --- |
|  | OR (95% CI) | OR (95% CI) | OR (95% CI) | OR (95% CI) | OR (95% CI) | OR (95% CI) | OR (95% CI) | OR (95% CI) |
| Age (years) | **1.02**  **[1.00, 1.04]** | **1.02**  **[1.00, 1.05]** | **1.02**  **[1.00, 1.04]** | **1.02**  **[1.00, 1.04]** | **1.02**  **[1.00, 1.04]** | **1.02**  **[1.00, 1.04]** | **1.02**  **[1.00, 1.04]** | **1.04**  **[1.01, 1.07]** |
| Height  (per 10 cm increase) | 1.18  [0.97, 1.44] | 1.19  [0.97, 1.46] | 1.18  [0.97, 1.44] | 1.19  [0.97, 1.45] | 1.21  [0.98, 1.49] | 1.19  [0.98, 1.46] | 1.18  [0.97, 1.44] | **1.39**  **[1.05, 1.83]** |
| Sex (Male) | 0.73  [0.50, 1.07] | 0.71  [0.48, 1.04] | 0.74  [0.51, 1.08] | 0.72  [0.49, 1.05] | 0.68  [0.46, 1.01] | 0.70  [0.48, 1.03] | 0.73  [0.50, 1.06] | 0.61  [0.35, 1.06] |
| Ever smoking  (vs never smoking) | 1.02  [0.68, 1.52] | 1.05  [0.69, 1.58] | 0.97  [0.64, 1.46] | 1.05  [0.70, 1.58] | 0.95  [0.62, 1.45] | 1.08  [0.72, 1.63] | 1.03  [0.69, 1.54] | 0.75  [0.44, 1.27] |
| Study site (Soweto) |  | | | | | | | |
| Agincourt | **1.78**  **[1.04, 2.99]** | 1.90  [0.89, 3.86] | **1.82**  **[1.07, 3.06]** | **1.79**  **[1.05, 3.02]** | 1.59  [0.85, 2.88] | **1.82**  **[1.07, 3.08]** | **1.81**  **[1.06, 3.04]** | 1.18  [0.61, 2.22] |
| DIMAMO | 1.38  [0.58, 2.91] | 1.00  [0.35, 2.44] | 1.38  [0.58, 2.93] | 1.39  [0.58, 2.95] | 1.36  [0.54, 2.99] | 1.52  [0.64, 3.24] | 1.43  [0.60, 3.03] | 1.33  [0.55, 2.85] |
| Nanoro | **4.27**  **[3.20, 5.73]** | 2.11  [0.71, 6.08] | **4.39**  **[3.28, 5.93]** | **4.49**  **[3.33, 6.10]** | **4.18**  **[3.12, 5.65]** | **5.18**  **[3.76, 7.19]** | **4.40**  **[3.28, 5.94]** | **4.16**  **[2.45, 7.00]** |
| SES quintile  (continuous score) | 0.82  [0.62, 1.07] | 0.82  [0.63, 1.08] | 0.82  [0.63, 1.08] | 0.80  [0.61, 1.06] | 0.83  [0.63, 1.10] | 0.83  [0.63, 1.09] | 0.82  [0.62, 1.08] | 0.76  [0.52, 1.11] |
| Solid fuel exposure (reference: no solid fuel use) |  | | | | | | | |
| Solid fuel use with  kitchen ventilation |  | 2.05  [0.73, 5.91] |  |  |  |  |  |  |
| Solid fuel use with  no kitchen ventilation |  | 2.01  [0.60, 6.84] |  |  |  |  |  |  |
| Prior TB (yes vs no) |  |  | 1.46  [0.84, 2.44] |  |  |  |  |  |
| Diabetes (yes vs no) |  |  |  | 1.38  [0.84, 2.22] |  |  |  |  |
| CVD (yes vs no) |  |  |  |  | 1.10  [0.68, 1.72] |  |  |  |
| Hypertension (yes vs no) |  |  |  |  |  | **1.46**  **[1.07, 2.00]** |  |  |
| CKD (yes vs no) |  |  |  |  |  |  | 1.45  [0.69, 2.81] |  |
| HIV infection  (yes vs no) |  |  |  |  |  |  |  | 1.38  [0.84, 2.21] |
| Overall N | 1619 | 1539 | 1617 | 1575 | 1530 | 1590 | 1613 | 1089 |
| *Note: Values are adjusted ORs (95% CIs). Base models included age, height, sex, ever smoking, study site and SES (continuous quintile score); each additional exposure was modelled separately, so N varies with data completeness. Bold denotes associations for which the 95% CI excluded the null value before rounding; some confidence limits round to 1.00. TB, tuberculosis; CVD, cardiovascular disease; CKD, chronic kidney disease; HIV, human immunodeficiency virus.* | | | | | | | | |
